# Stability of Extemporaneously Prepared Preservative-Free Methylphenidate 5 mg/mL Intravenous Solution

**DOI:** 10.1101/2020.08.29.20184457

**Authors:** Megan E. Barra, Brian L. Edlow, James T. Lund, Katherine Sencion, John Vetrano, Cherylann Reilly-Tremblay, Edlyn R. Zhang, Yelena Bodien, Emery N. Brown, Ken Solt

## Abstract

**Purpose:** To advance the implementation of consciousness-promoting therapies in patients with acute disorders of consciousness, the availability of potential therapeutic agents in formulations suitable for administration in hospitalized patients in the presence of complex comorbid conditions is paramount. The purpose of this study is to evaluate the long-term stability of extemporaneously prepared preservative-free methylphenidate 5 mg/mL intravenous solution.

**Methods:** A methylphenidate 5 mg/mL solution was prepared under proper aseptic techniques with Methylphenidate HCl USP powder mixed in sterile water for solution. Methylphenidate HCl 5 mg/ml solution was sterilized by filtration technique under USP <797> compliant conditions. Samples were stored refrigerated (2 - 8°C) and analyzed at approximately day 1, 30, 60, 90, 180, and 365. At each time point, chemical and physical stability was evaluated by visual inspection, pH measurement, membrane filtration procedure, turbidometric or photometric technique, and high-performance liquid chromatography (HPLC) analysis.

**Results:** Over the 1-year study period, the sample retained 96.76 – 102.04% of initial methylphenidate concentration. There was no significant change in the visual appearance, pH level or particulate matter during the study period. The sample maintained bacterial and fungi sterility, and endotoxin levels were undetectable throughout the 1-year stability period.

**Conclusions:** Extemporaneously prepared preservative-free methylphenidate 5 mg/mL intravenous solution was physically and chemically stable for up to 365 days when stored in amber glass vials at refrigerated temperatures (2 - 8°C).

## Introduction

Methylphenidate hydrochloride (HCl) is classified as a Schedule II controlled substance by the United States (U.S.) Drug Enforcement Administration and approved by the U.S. Food and Drug Administration for the treatment of attention-deficit/hyperactivity disorder.^1,2^ Methylphenidate has been used off-label for the management of major depressive disorder, narcolepsy, severe fatigue in the cancer or palliative care setting and is increasingly utilized as a neurostimulant to promote recovery after traumatic brain injury, cardiac arrest, and stroke in the acute and chronic recovery periods.^3^–^8^ It is actively being explored as a therapeutic agent to promote emergence of consciousness after generalized anesthesia or in patients with severe traumatic brain injury.^9^–^11^ With the Neurocritical Care Society Curing Coma Campaign launch in 2019 to strategically advance the implementation of consciousness-promoting therapies in patients with disorders of consciousness, the availability of potential therapeutic agents in formulations suitable for administration in hospitalized patients in the presence of complex comorbid conditions is paramount.^12^

Methylphenidate is currently available in the U.S. as immediate release, sustained release, and extended-release oral suspension, chewable, and disintegrating tablets and/or capsules, transdermal patch, and bulk powder.^13^ Methylphenidate HCl is supplied as a racemic mixture dl-threo-methylphenidate subject to enantioselective first-pass metabolism.^2^ Intravenous methylphenidate may be advantageous over enteral administration due to its more predictable absorption profile.^13^–^15^ Additionally, intravenous methylphenidate may be administered to hospitalized patients who are contraindicated for enteral administration due to post-operative status, gastrointestinal dysfunction during hospitalization, or for acute treatment to promote recovery of consciousness in the research setting.^10,11^ In the absence of stability and sterility data, the U.S. Pharmacopeia (USP) beyond-use date of this non-sterile to sterile compounded solution is limited to 72-hours refrigerated or 24-hours room temperature.^16^

The purpose of this study was to evaluate the long-term stability of extemporaneously prepared preservative-free methylphenidate 5 mg/mL intravenous solution.

## Materials and Methods

### Solution Preparation

Compounded solution was prepared by the Department of Pharmacy at Massachusetts General Hospital in Boston, Massachusetts. An intravenous stock solution of methylphenidate 5 mg/mL was prepared in a glass beaker by dissolving 1950 mg methylphenidate hydrochloride USP powder (Letco Medical) in sterile water for injection to a final volume of 390 mL. Mixed solution pH was adjusted to range pH 3-4 using hydrochloric acid or sodium hydroxide. Under a laminar flow hood under USP <797> aseptic conditions, non-sterile methylphenidate 5 mg/mL 390 mL solution was filtered into an empty polyvinyl chloride plastic container using a 0.22 micron filter (Sterivex-GP pressure filter unit) to sterilize the solution. A total filtered solution volume of 15.2 mL was packaged into each 30 mL amber glass vial. Amber glass vials were sealed using tamper evident seals and labeled accordingly. Vials were stored under refrigerated conditions (2-8°C) and transported to a third-party lab for subsequent testing. Forty-six vials were transported for validation testing followed by 25 vials for stability testing approximately 1 month later. Refrigerated samples were stored in a dark refrigerator and exposed to light only during sampling. These samples were allowed to warm at room temperature without application of external heat sources.

Physical and chemical stability of the methylphenidate 5 mg/mL 15 mL solution was evaluated at approximately 1, 30, 60, 90, 180, and 365 days. Physical and chemical stability was performed by DynaLabs LLC (St. Louis, Missouri). Chemical stability was evaluated using high-performance liquid chromatography (HPLC) as outlined in USP <621>. Microbial growth for bacteria and fungi was evaluated using membrane filtration procedures pursuant to USP <71>. Particulate matter was examined using a light obscuration particle count test in accordance with USP <788>. Visual inspection for clarity and color as specified in USP <797> was additionally examined at these time-points.

### Chromatographic Apparatus and Conditions

Chromatographic studies were performed using a Phenomenex Gemini C18 HPLC column. The mobile phase was a 65/35 ratio of 2.7 g/L solution of potassium phosphate monobasic in water buffer and methanol. The flow rate was 1.8 mL/min for 0.5 min and 10 min for Step 0 and Step 1, respectively.

### Validation of Methylphenidate Hydrochloride HPLC Method

Calibration curves were developed using linear regression of the calculated areas versus theoretical concentration (µg/mL). Acceptance criterion was set at r^2^ > 0.999. Accuracy was determined using a standard stock solution of 10 mg Methylphenidate Hydrochloride USP standard powder diluted with potassium phosphate monobasic in water 2.7 g/L solution (Fisher Scientific) to a final concentration of 500 µg/mL. Samples were further diluted according to a dilution schedule to a concentration 15 µg/mL (n=3), 25 µg/mL (n=3), or 35 µg/mL (n=3). Acceptance criterion for accuracy of average % recovery was set at > 97.5% and < 102.5%. Precision was determined using the same standard stock solution formula as the accuracy methods. Samples were diluted according to a dilution schedule to 25 µg/mL (n=9). Percent relative standard deviation (RSD) for the retention time and areas was calculated and acceptance criteria was set at < 2% RSD. Precision analysis was repeated on a different day and different instrument to determine intermediate precision (ruggedness).

### Forced Degradation Study

A forced degradation study was performed to ensure the HPLC method was indicative of stability and capable of separation of degradation products from parent drugs. Forced degradation stock solution was compounded using 4.5 mg Methylphenidate HCl diluted with potassium phosphate monobasic (Fisher Scientific) in water 2.7 g/L solution to a final concentration of methylphenidate 90 µg/mL. Triplicate samples were prepared in 2-dram amber vials except for light samples, which were prepared in clear 2-dram vials, and diluted to 30 µg/mL. Samples underwent either acid hydrolysis, base hydrolysis, or oxidization using 1 mL of 0.1M hydrochloric acid solution (Fisher Scientific), sodium hydroxide (Fisher Scientific), or 3.5% H^2^O^2^ (Honeywell HPLC grade), respectively. Light samples were placed 8 cm apart from a 365 nm UV lamp light and left static for 3 hours, after which 1-mL of water was added to the sample and placed into amber liquid chromatography (LC) vials. Heat samples were placed in a beaker filled with deionized water heated to 70 °C on top of a heated stir plate for 3-hours, after which vials were removed and allowed to cool to room temperature, mixed with 1 mL of water (Honeywell HPLC grade) and placed into amber LC vials. Liquid chromatography sequencing was performed on all standard forced degradation samples after an initial blank sample was run. The area under three standard injections was averaged to one value. For all other samples, the percentage recovery was determined by dividing the areas of the samples by the average area of the standard and multiplying by 100. Purity values were analyzed according to peak apex and similarity algorithm for 5-distinct portions. Values > 900 were considered pure while values < 900 were considered pure only if value is within 10% of the standard values.

### Data Analysis

The stability of methylphenidate 5 mg/mL intravenous solution was determined. Stability was defined as at least 90% retention of the initial Methylphenidate HCl concentration at each time-point evaluated.

## Results and Discussion

At least 95% of the initial concentration of methylphenidate was retained throughout the 1-year study period (Table 1). No detectable changes upon visual inspection in color or clarity were observed.

**Table 1.** Stability of Methylphenidate Hydrochloride 5 mg/mL Intravenous Solution Stored at 2 - 8°C.

| *Methylphenidate HCl* |  |  |
| --- | --- | --- |
| *Day* | **Result** | **% Initial Concentration Remaining** |
| 1 | 5.10 mg/mL | 102.04% |
| 32 | 4.96 mg/mL | 99.14% |
| 61 | 5.06 mg/mL | 101.15% |
| 95 | 5.03 mg/mL | 100.52% |
| 186 | 4.84 mg/mL | 96.76% |
| 365 | 4.93 mg/mL | 98.68% |

Furthermore, no microbial growth was detected at any point during the 1-year study period and endotoxin were non-detectable < 0.05 EU/mL. No appreciable change in pH (range 3.43-3.67) over the 1-year study period was observed.

The number of particles > 10 µm per container were 102 parts/container on day 4, 47 parts/container (day 33), 70 parts/container (day 68), 52 parts/container (day 95), 28 parts/container (day 182), and 155 parts/container (day 375). The number of particles > 25 µm per container were 0 parts/container on day 4, 0 parts/container (day 33), 4 parts/container (day 68), 2 parts/container (day 95), 2 parts/container (day 182), and 6 parts/container (day 375).

## Conclusions

Extemporaneously prepared preservative-free methylphenidate 5 mg/mL intravenous solution was stable for up to 365 days when stored refrigerated in 30-mL amber glass bottles.

## Data Availability

The data referred to in this manuscript is available upon request.

## Financial Disclosures

The study was funded by NIH Director’s Office (DP2HD101400). The authors declare no relevant conflicts of interest.

## Notes

### Competing Interest Statement

The authors have declared no competing interest.

### Funding Statement

The study was funded by NIH Directors Office (DP2HD101400).

### Author Declarations

This study was exempt by the Massachusetts General Hospital Institutional Review Board.

